# THE EFFECT OF THE MEASLES, MUMPS AND RUBELLA VACCINE ON INNATE AND ADAPTIVE IMMUNE RESPONSES IN PERSONS RECEIVING A SARS-COV-2 mRNA VACCINE

**DOI:** 10.1101/2022.09.09.22279771

**Authors:** Leon du Toit, Ananya Gupta, Hakim-Moulay Dehbi, Tamarand L. Darling, Darya V. Urusova, Shyamala Thirunavukkarasu, Adrianus C.M. Boon, Erik R. Dubberke, Linda Yun, Sherry McKinnon, Anne K. DeSchryver, Ben Swan, Mihai G. Netea, Shabaana Khader, Michael S. Avidan, the CROWN CORONATION Investigators

**Affiliations:** Department of Anesthesia, Washington University School of Medicine, Saint Louis, MO, USA; Department of Anaesthesia and Perioperative Medicine, University of Cape Town, South Africa; Department of Molecular Microbiology, Washington University School of Medicine, Saint Louis, MO, USA; Comprehensive Clinical Trials Unit, Institute of Clinical Trials and Methodology, UCL, London, UK; Department of Medicine, Washington University School of Medicine, Saint Louis, MO, USA; Department of Pathology and Immunology, Washington University School of Medicine, Saint Louis, MO, USA; Division of Infectious Diseases, Washington University School of Medicine, Saint Louis, MO, USA; Department of Internal Medicine and Radboud Center for Infectious Diseases, Radboud University Medical Centre, Nijmegen, The Netherlands; Department of Immunology and Metabolism, Life & Medical Sciences Institute, University of Bonn, Germany

## Abstract

Vaccination elicits a complex combination of immune responses. Immune memory formation is observed not only in the antibody responses of B-cells, but also in the T-cell response. Moreover, some live attenuated vaccines such as measles-containing vaccines can induces heterologous protection, likely through induction of memory characteristics in the innate immune response. Little is known about the immunological interaction that may occur when different vaccines are administered soon after one another, especially in relation to the novel COVID-19 vaccines.

The aim of this study was to compare the innate and adaptive immune responses between persons randomized to receive either a MMR or a placebo (0.9% NaCl) injection prior to their SARS-CoV-2 mRNA vaccination. We compared: i) the cytokine and chemokine production (tumor necrosis factor [TNF]-α, interleukin [IL]-1β, IL-6, IL-10, IL-17, IL-22, interferon [IFN]-α and IFN-γ) after in-vitro stimulation of peripheral blood mononuclear cells (PBMCs) with heterologous stimuli (severe acute respiratory syndrome coronavirus [SARS-CoV]-2, measles mumps and rubella [MMR] vaccine, Toll-like receptor [TLR]-3 ligand, TLR-7/8 ligand, or TLR-4 ligand), and ii) the SARS-CoV-2 neutralizing antibody responses.

Ninety-five participants in the CROWN CORONATION trial (NCT04333732; a randomized control trial comparing MMR to placebo for prevention of COVID-19) agreed to an additional single blood sample collection for this immunological study. Samples were collected around 196 (SD 22) days after administration of MMR or placebo, and around 105 (SD 27) days after their second SARS-CoV-2 mRNA vaccine injection.

Twenty-four percent of participants were older than fifty and sixty-seven percent were female. The median TNF-α response to stimulation with MMR was 8315.3 pg/mL in the MMR group and 4340.5 pg/mL in the placebo group; adjusted median difference (95% CI) 3012.5 (−4734.1; −323.5); p=0.017. No other significant differences were noted in the cytokine and chemokine responses between treatment groups. The SARS-CoV-2 neutralization assay geometric mean (SD) IC50 in the MMR group was 507.6 (2.6) and in the placebo group was 515.7 (2.2); ratio of geometric means (95% CI) 1.0 (0.7; 1.5).

Pre-exposure to MMR vaccine was generally not associated with changes in cytokine and chemokine responses of stimulated PBMCs at 105 (27) days after SARS-CoV-2 mRNA vaccination. MMR vaccination led only to an increase of TNF-α production in response to an additional ex-vivo stimulation with the MMR vaccine. The SARS-CoV-2 neutralization IC50 values did not differ between MMR and placebo groups. Further studies using a repeated measures design would be better suited to explore or rule-out any short-lived vaccine response and vaccine-vaccine immunological interaction.

## Background

The production of neutralizing antibodies (especially IgG) by B-cells underpins our most basic understanding of vaccine-derived immunity and is usually the correlate of protection used for assessing individual vaccine response and immunity in a clinical setting (Ada, 2001; Iwasaki and Omer, 2020). Specific memory against a pathogen is also built by T cells, which amplify the function of other immune cells and can eliminate virus-infected cells. Moreover, it is now recognized that the innate immune system plays a key role in the vaccine response and innate immune cells themselves have memory-like responses (also termed ‘*trained immunity’*) that can enhance resistance to heterologous infection (Domínguez-Andrés et al., 2019). Trained immunity is mediated by the functional reprogramming of innate immune cells such as monocytes, macrophages, and Natural Killer (NK) cells, resulting in lasting enhanced responsiveness of these cells to antigen-independent heterologous stimuli (Netea et al., 2020).

Epidemiological data demonstrate heterologous protective effects of certain vaccines against infections other than those for which these vaccines are administered (Higgins et al., 2016). This off-target protection cannot be explained by induction of antigen-specific T- or B-cell responses, but may instead be explained by either heterologous T-cell responses or trained immunity (Benn et al., 2013). Vaccination with a live attenuated measles virus induces lymphoproliferative responses (Bautista-López et al., 2000), but whether it also induces an increase in innate immune responses, as measured by heterologous cytokine production, has not yet been studied.

The immune response to a vaccine encompasses an ensemble of responses of variable duration by multiple components of the immune system. The question arises on how does recent exposure to a vaccine affect the immune response to subsequent vaccination with a second unrelated vaccine? Such immunological interactions between different vaccines have not been studied extensively (Gizurarson, 1998), and evidence suggests that the chronological order of different vaccines in childhood immunization schedules can have a notable impact on overall childhood mortality (Shann, 2021).

The rapid development and roll-out of specific SARS-CoV-2 mRNA vaccination in the USA at the end of 2020 coincided with enrolment of participants into the CROWN CORONATION trial (NCT04333732) – a study investigating the efficacy of the measles, mumps and rubella (MMR) vaccine to prevent symptomatic, PCR positive COVID-19. Subsequent to randomization to either MMR or placebo injection, the majority of CROWN CORONATION trial participants in the USA were vaccinated with a SARS-CoV-2 mRNA vaccine. These events created the unique opportunity to observe in a randomized population the effect that MMR vaccination has on innate immune responses and on the immune responses induced by subsequent administration of a specific SARS-CoV-2 vaccine.

The aim of this sub-study is to characterize, in those pre-exposed to a recent MMR or placebo vaccination followed by mRNA SARS-CoV-2 vaccine administration, the comparative in-vitro cytokine and chemokine responses to heterologous stimuli and the SARS-CoV-2 neutralizing antibody responses.

We hypothesize that, compared to SARS-CoV-2 mRNA vaccination alone, MMR vaccination prior to SARS-CoV-2 mRNA vaccination will induce lasting trained immunity in recipients, manifested by a stronger innate immune response upon in vitro exposure of supernatants to heterologous pathogen products. We also hypothesize that MMR vaccination prior to SARS-CoV-2 mRNA vaccination does not alter the adaptive immune response to SARS-CoV-2 mRNA vaccination.

## Methods

The hypotheses were tested in a sub-study within the CROWN CORONATION randomized control trial. Sub-study participation required written informed consent for an additional study visit and blood sample collection at the Washington University in St Louis study site. The sub-study protocol was approved by the biomedical IRB at Washington University in St Louis (IRB ID: 202011081) and registered on the clinicaltrials.gov registry (NCT04646239) prior to recruitment.

### Participant enrolment and study design

To take part in the sub-study, a prospective participant had to be enrolled in the CROWN CORONATION trial and have received two doses of a SARS-CoV-2 mRNA vaccine after receiving their investigational medicinal product (IMP) injection of either MMR or 0.9% NaCl placebo. The study population was contacted telephonically to elicit enrolment into the sub-study.

Participants and the study personnel involved in recruitment, the consent process, and laboratory personnel were blinded to the group allocation of the participants. The research nurse who collected the samples and the statistician were the only unblinded study team members.

The primary outcomes recorded in the sub-study were peripheral blood mononuclear cell (PBMC) cytokine and chemokine production (tumor necrosis factor [TNF]-α, interleukin [IL]-1β, IL-6, IL-10, IL-17, IL-22, interferon [IFN]-α and IFN-γ) after in vitro stimulation with heterologous stimuli (inactivated SARS-CoV-2, live attenuated MMR viruses, Toll-like receptor [TLR]-3 ligand, TLR-7/8 ligand, TLR-4 ligand, and Roswell Park Memorial Institute [RPMI] medium; Table 1). The secondary outcomes were SARS-CoV-2 neutralization assay (IC50) and measles IgG antibody titres.

**Table 1:** Ex vivo stimulation of peripheral blood mononuclear cells (PBMCs)

| # | Stimulus | Concentration |
| --- | --- | --- |
| S1 | Roswell Park Memorial Institute (RPMI) medium |  |
| S2 | TLR-4 ligand: LPS | 10 ng/ml |
| S3 | TLR-3 ligand: Poly(I:C) | 10 μg/ml |
| S4 | TLR-7/8 ligand: R848 | 3 μg/ml |
| S5 | MMR2 | 4.9x10<sup>3</sup> PFU/ml |
| S6 | SARS-CoV-2 | 1.4x10<sup>3</sup> PFU/ml |

### Sample collection

Blood collection took place at a single study visit around 12 to 18 weeks following the participant’s second SARS-CoV-2 mRNA vaccine injection. 45 ml of blood was collected from each sub-study participant. Blood was collected in K2EDTA tubes for PBMC stimulation testing and SARS-CoV-2 neutralization assay and in serum separator tubes for measurement of measles IgG antibody titres. Collected specimens were transferred to the research laboratories within two hours of collection.

### PBMC isolation from blood

All downstream processing was carried out aseptically. Briefly, the tubes were centrifuged (3800 *rpm*; 10 minutes at room temperature) to separate plasma and cellular phase of the blood samples collected. The top layer of plasma was collected in sterile tubes and stored at −80^0^ C freezer for future analysis (including SARS-CoV-2 neutralization assay). The cellular phase was used for PBMC isolation. The cellular fraction of blood in K2EDTA tubes was transferred to 50 mL conical tubes (BD Falcon). Sample collection tubes were rinsed with ice cold PBS (supplemented with Heparin and 2% human serum) to recover any residual blood into the conical tube. The volume of blood per conical tube was adjusted to 35-40 mL using PBS. For density gradient-based separation of PBMC from blood, 10 mL of Ficoll was slowly added to a fresh 50 mL conical tube using a sterile pipette. Blood was layered on Ficoll and centrifuged for density gradient separation at 1700 rpm at 4°C for 30 minutes without break (i.e., Deceleration at 0 and Acceleration at 2). The interphase so obtained was collected separately, washed twice with non-supplemented RPMI-1640 and pellet was resuspended in 5 mL of cRPMI (RPMI-1640 supplemented with 10% human serum, 1% Penicillin-Streptomycin and 1% L-Glutamine) and cells were counted using haemocytometer. PBMCs were resuspended at a concentration of 5 million cells/mL.

### Ex-vivo PBMC stimulation

Isolated PBMCs were exposed to the heterologous stimuli (Table 1). 100 μL of the PBMC cell suspension (500,000 cells) per well were added to a 96-well flat bottom plate (NUNC, tissue culture grade). 100 μL of stimuli prepared in cRPMI were added to respective PBMC wells; control wells only received cRPMI. Plates were incubated at 37°C, 5% CO2 for 24 hours or 5 days, following which the supernatant was frozen at −80°C for downstream cytokine analysis.

### Cytokine and chemokine measurements

Supernatants stored at −80°C were subjected to cytokine measurements using a multiplex assay kit (Human cytokine/chemokine multiplex assay; Millipore Sigma) as per manufacturer’s protocol and were done in batches. TNF-α, IL-1β, IL-6, and IFN-α was measured in supernatants obtained 24 hours post stimulation and IL-10, IL-17, IL-22, and IFN-γ was measured in supernatants obtained five days post stimulation of PBMCs. All reagents were warmed to room temperature prior to the assay. The control and samples were incubated with equal volume of the bead mixture on a microplate shaker, overnight at 4°C in the dark. The wells were washed using a handheld magnetic plate separator followed by addition and incubation with detection antibody and streptavidin-phycoerythrin, respectively. Data was acquired on Bio-rad Bioplex200 Luminex machine using the Bio-plex Manager 6.1 software.

### Focus reduction neutralization titer assay (FRNT)

Serial dilutions of the human serum samples were incubated with 100 focus-forming units (FFU) of the WA1/2020 strains of SARS-CoV-2 for 1 h at 37°C. Antibody-virus complexes were added to Vero-cell monolayers in 96-well plates and incubated at 37°C for one hour. Subsequently, cells were overlaid with 1% (w/v) methylcellulose in Eagle’s Minimal Essential medium (MEM, Thermo Fisher Scientific) supplemented with 2% FBS. Plates were harvested 30 hours later by removing overlays and the cells were fixed with 4% paraformaldehyde (PFA) in PBS for 20 min at room temperature. Plates were washed and sequentially incubated with an oligoclonal pool of SARS-CoV-2 specific monoclonal antibodies (SARS2-2, SARS2-11, SARS2-16, SARS2-31, SARS2-38, SARS2-57, and SARS2-71) and HRP-conjugated goat anti-mouse IgG (Sigma Cat # A8924) in PBS supplemented with 0.1% saponin and 2% FBS. SARS-CoV-2-infected cell foci were visualized using TrueBlue peroxidase substrate (KPL) and quantitated on an ImmunoSpot microanalyzer (Cellular Technologies). The number of foci per well was normalized to control wells and the serum dilution inhibiting foci formation by 50% (IC50) was calculated in GraphPad Prism 9.3.

### Sample size

We aimed to enroll as many participants as possible into the sub-study. The eligible population size was 145 CROWN CORONATION trial participants at the Washington University in St Louis study site who remained in the trial and had received two doses of an mRNA SARS-CoV-2 vaccine. The allocation ratio in the main trial was 1:1 to MMR or 0.9% NaCl. To predict the power of our study, we used the t-test to compare the primary outcome between treatment arms, in other words, the difference in cytokine response between MMR and placebo groups. Table 2 reports power given the expected difference in IFN-γ responses, with a two-sided type-1 error rate of 5%. The expected values were obtained from unpublished data on the PBMC response to stimulation with Bacillus Calmette-Guérin (BCG) in a separate study by one of our group (MN).

**Table 2:** Predicted power.

| Stimulus | Expected difference between means; IFN-γ (pg/ml) | Expected standard deviation | Power with 60 participants | Power with 100 participants | Power with 140 participants |
| --- | --- | --- | --- | --- | --- |
| TLR-3 ligand | 400 | 900 | 39% | 59% | 74% |
| TLR-7/8 ligand | 450 | 900 | 48% | 70% | 84% |

### Statistical analysis

This sub-study is hypothesis generating. No adjustment was made for multiple testing. We used complete case analysis throughout. All endpoint analyses were according to Intention-To-Treat (ITT). Baseline characteristics are summarised as mean and standard deviation for continuous (approximate) normally distributed variables, medians and interquartile ranges for non-normally distributed continuous variables, and frequencies and percentages for categorical variables. We used histograms of the continuous variables to assess normality. Endpoints are summarised graphically with box plots and dot plots.

Primary endpoint analysis was by median regression, with a coefficient for treatment arm, and adjusted for sex and the calendar date (quarter of the year) of sample collection. These adjustments were prespecified because sex (Aaby et al., 2020) and environmental allergen exposure (Gilles et al., 2020) which correlates with calendar date within a location are known determinants of an individual’s immune response (Saint Louis County Department of Health, 2021). The endpoints are reported as the adjusted median (95% CI) effect of MMR pre-exposure on the endpoint. For each participant the IL-1β response to RPMI was evaluated and if the response was greater than 100 pg/ml, the sample was considered to be contaminated and the data from that participant was excluded from analysis for all primary endpoints. Difference in virus neutralization (IC50) between treatment groups were analysed with multiple linear regression, adjusted for sex and calendar date. IC50 values were log(e) transformed and findings are reported as unadjusted geometric means and the ratio of geometric means adjusted for sex and quarter of the year. Difference in measles IgG titres between treatment groups were analysed with a simple logistic regression. IgG titres ≥ 16.5 AU/ml are considered consistent with immunity against measles according to the reference laboratory. The lower and upper limits of detection of the assay are 13.5 AU/ml and 300.0 AU/ml.

Sensitivity analysis was conducted excluding participants who were diagnosed with COVID-19 during the trial period prior to their blood draw.

All analyses were conducted using Rstudio (version 1.4.1717, “Juliet Rose”, Boston, MA).

## Results

### Effects of MMR vaccination on cytokine production capacity

From 22 March 2021 to 28 June 2021, 95 participants (representing 66% of the eligible population) were recruited and presented for blood draws at the study site (Figure 1).

**Figure 1:**
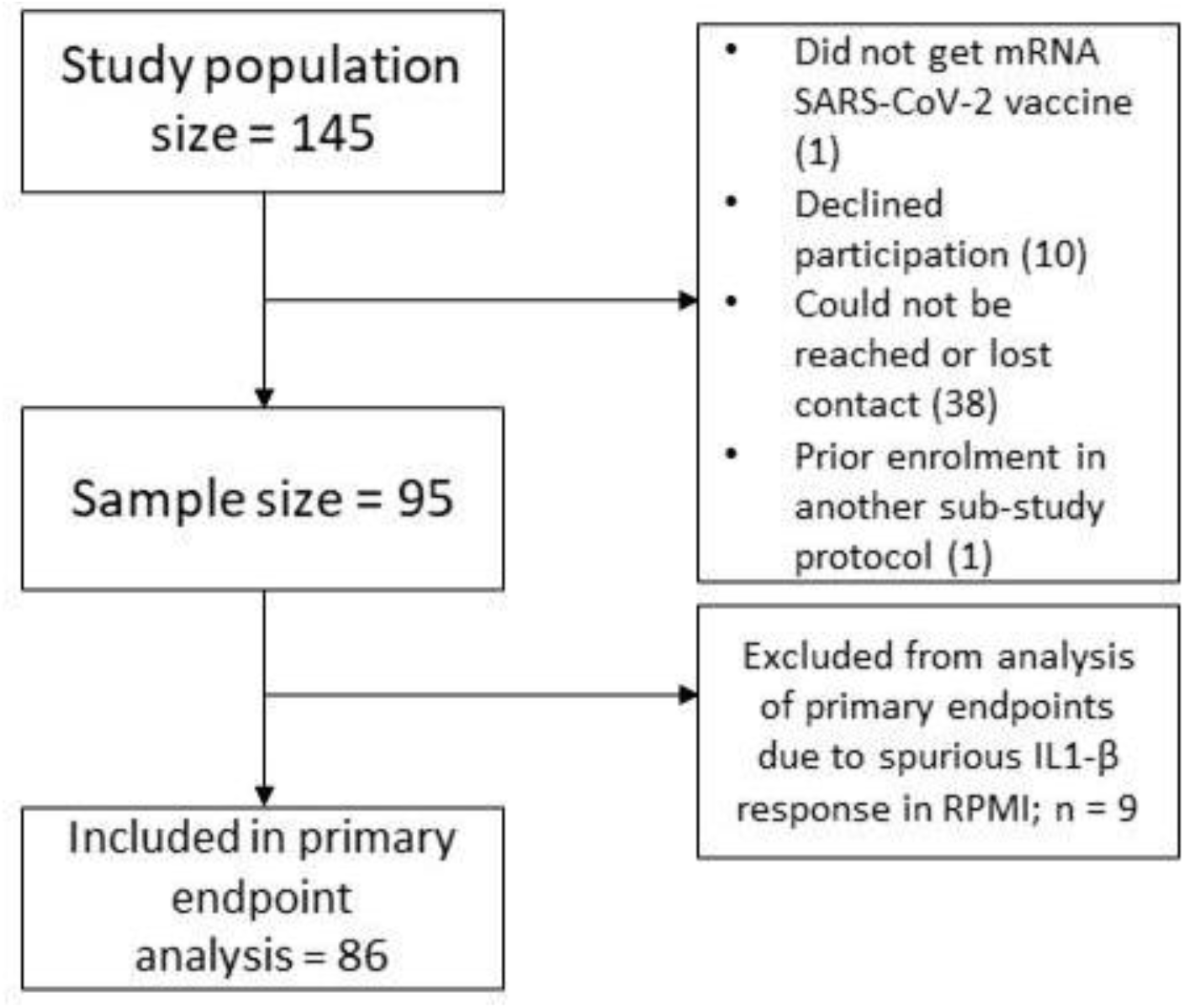
Participant flow diagram

Forty-eight participants were exposed to MMR and 47 were exposed to placebo prior to their mRNA SARS-CoV-2 vaccination. Measured baseline characteristics appear balanced between the MMR and placebo groups; Table 3. Twenty-three (24.2%) participants were older than 50 years of age. Sixty-four (67.4%) were female. On average (sd), blood sampling took place 196 (22) days after IMP administration (MMR or 0.9% NaCl) and 105 (27) days after the last SARS-CoV-2 mRNA vaccine injection. Participants received their first dose of SARS-CoV-2 mRNA vaccine 71 (26) days after injection of the IMP. Six of the study participants were diagnosed with symptomatic COVID-19 prior to their blood draws. All six events occurred prior to their first SARS-CoV-2 mRNA vaccination.

**Table 3:**
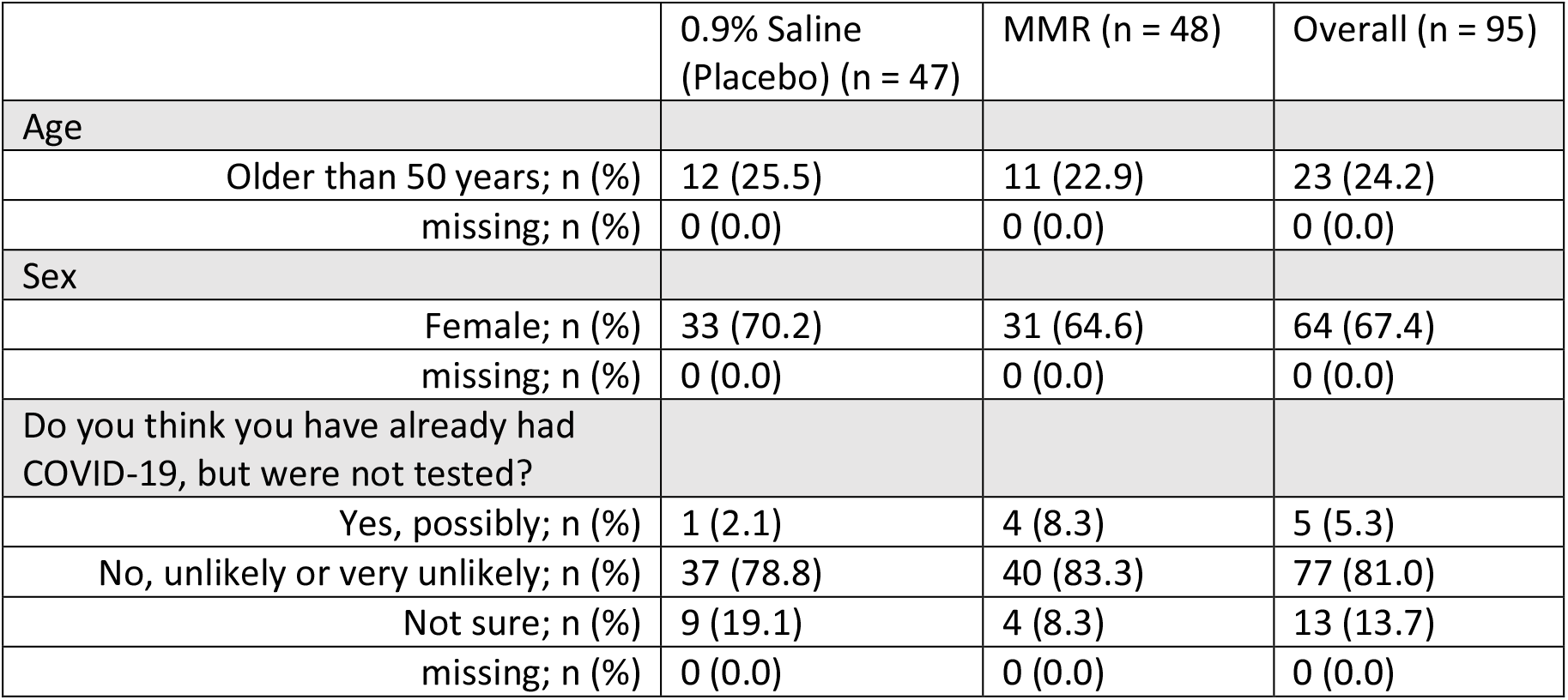
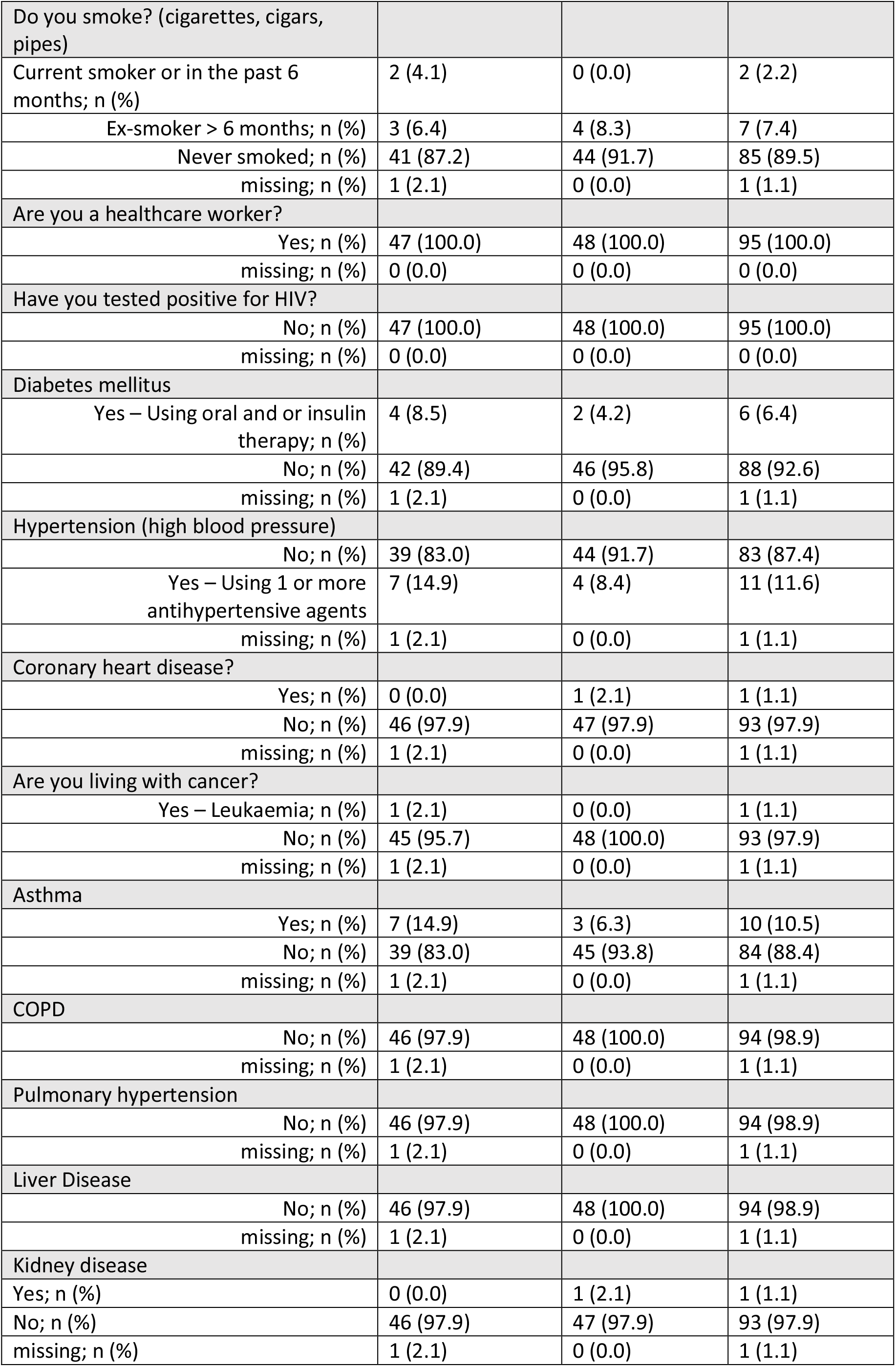
Baseline characteristics reported as mean (sd), or median [lower quartile, upper quartile] or count (%)

The PBMC cytokine and chemokine responses following the stimuli are summarised in Figures 2-6. IL-17, IL-22 and IFN-α production was not induced by any of the stimuli and therefore not specifically depicted. Table 4 reports the chemokine and cytokine responses adjusted for sex and period of the year. The IL-6 responses to LPS (TLR4 ligand), Poly(I:C) (TLR3 ligand), R848 (TLR7/8 ligand) and to MMR appear truncated by the upper limit of the assay and are therefore not specifically reported. The median TNF-α response to stimulation with MMR was 8315.3 pg/mL in the MMR group and 4340.5 pg/mL in the placebo group; adjusted median difference (95% CI) 3012.5 (−4734.1; −323.5); p=0.017. There was no notable difference between the treatment groups for any of the other cytokine or chemokine responses to any of the heterologous stimuli.

**Table 4:** Peripheral blood mononuclear cell (PBMC) chemokine and cytokine responses to heterologous stimuli. Median differences adjusted for sex and quarter of the year.

|  | MMR median response; pg/ml; N = 46 | Placebo median response; pg/ml; N = 40 | Adjusted difference between medians (95% CI); pg/ml | p-value |
| --- | --- | --- | --- | --- |
| **TNF-α** |  |  |  |  |
| RPMI (control) | 13.0 | 10.3 | -2.5 (-8.6; 2.9) | 0.606 |
| **MMR** | **8315.3** | **4340.5** | **-3012.5 (-4734.1; -323.5)** | **0.017\*** |
| SarsCoV2 | 20.0 | 14.3 | -2.0 (-14.3; 7.6) | 0.840 |
| Poly(I:C) (TLR3 ligand) | 1965.0 | 1456.3 | -282.5 (-825.7; 223.1) | 0.245 |
| R848 (TLR7/8 ligand) | 8739.0 | 8491.5 | -242.5 (-2040.9; 2344.9) | 0.877 |
| LPS (TLR4 ligand) | 4745.5 | 3291.5 | -781.0 (-2222.0; -160.4) | 0.289 |
| **IL-1β** |  |  |  |  |
| RPMI (control) | 0.5 | 0.5 | 0.0 (0.0; 0.0) | 1.000 |
| MMR | 8902.8 | 8864.5 | 566.5 (-2296.1; 2733.5) | 0.727 |
| SarsCoV2 | 1.5 | 2.5 | 0.0 (-2.7; 8.0) | 0.997 |
| Poly(I:C) (TLR3 ligand) | 3825.5 | 3985.8 | 460.0 (-1368.5; 1620.5) | 0.718 |
| R848 (TLR7/8 ligand) | 11806.5 | 12370.3 | 865.5 (-593.5; 2610.7) | 0.331 |
| LPS (TLR4 ligand) | 10723.0 | 11353.0 | 1083.3 (-599.8; 2340.2) | 0.410 |
| **IL-6** |  |  |  |  |
| RPMI (control) | 707.0 | 644.5 | -41.0 (-233.9; 121.1) | 0.753 |
| SarsCoV2 | 854.5 | 769.0 | -53.0 (-223.2; 19.8) | 0.456 |
| **IL-10** |  |  |  |  |
| RPMI (control) | 141.0 | 166.5 | 5.5 (-64.6; 50.8) | 0.871 |
| MMR | 5005.8 | 4216.0 | -646.5 (-2233.9; 89.4) | 0.250 |
| SarsCoV2 | 136.0 | 153.0 | 17.0 (-31.7; 43.7) | 0.491 |
| Poly(I:C) (TLR3 ligand) | 3651.8 | 3506.0 | -153.0 (-1126.3; 746.0) | 0.823 |
| R848 (TLR7/8 ligand) | 9594.0 | 9190.8 | 756.5 (-272.8; 1606.6) | 0.291 |
| LPS (TLR4 ligand) | 9515.5 | 9687.0 | 438.5 (-1285.4; 1098.7) | 0.583 |
| **IFN-γ** |  |  |  |  |
| RPMI (control) | 2.0 | 2.0 | 0.0 (-2.2; 1.4) | 1.000 |
| MMR | 48.3 | 51.5 | 3.0 (-8.1; 14.7) | 0.621 |
| SarsCoV2 | 1.8 | 2.0 | 0.5 (-1.6; 1.4) | 0.739 (b) |
| Poly(I:C) (TLR3 ligand) | 64.0 | 57.0 | -0.8 (-2.4; 1.9) | 0.604 |
| R848 (TLR7/8 ligand) | 1305.0 | 963.8 | 89.5 (-905.3; 540.1) | 0.823 |
| LPS (TLR4 ligand) | 113.3 | 94.0 | -20.0 (-54.7; -6.3) | 0.217 |

95% CIs computed by the rank inversion method (Koenker R, *Quantile Regression*, Section 3.4.5, Cambridge University Press, 2005). P-values assume iid, except where this method failed in which case bootstrap (b) method was applied. Lower detection limits for the cytokines in each assay as per manufacturer's protocol are as follows: TNF-α - 5.39 pg/ml; IL-1β - 0.52 pg/ml; IL-6 - 0.14 pg/ml; IL-10 - 0.91 pg/ml; IFN-γ - 0.86 pg/ml

**Figure 2:**
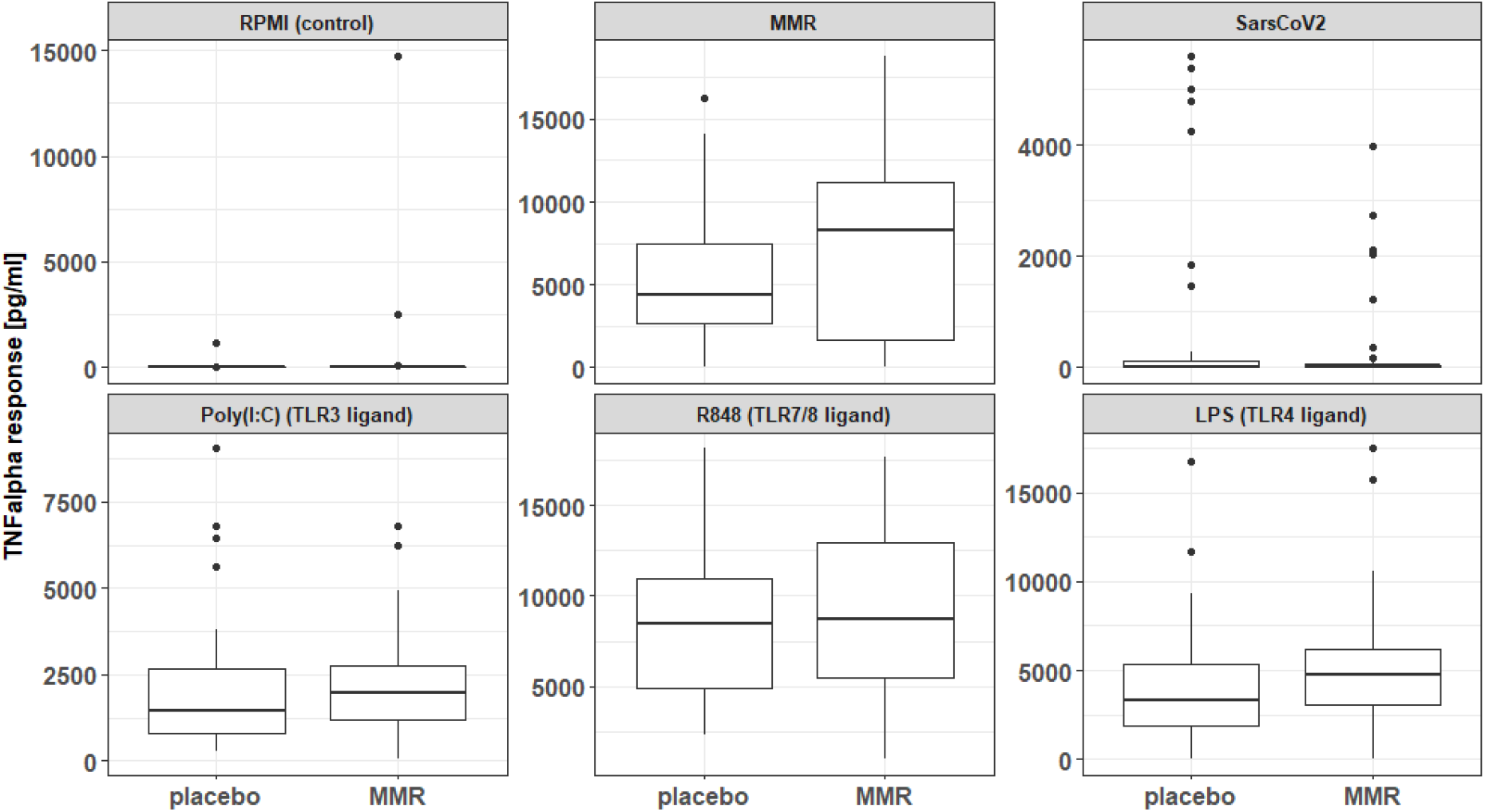
TNF-α responses to heterologous stimulation

**Figure 3:**
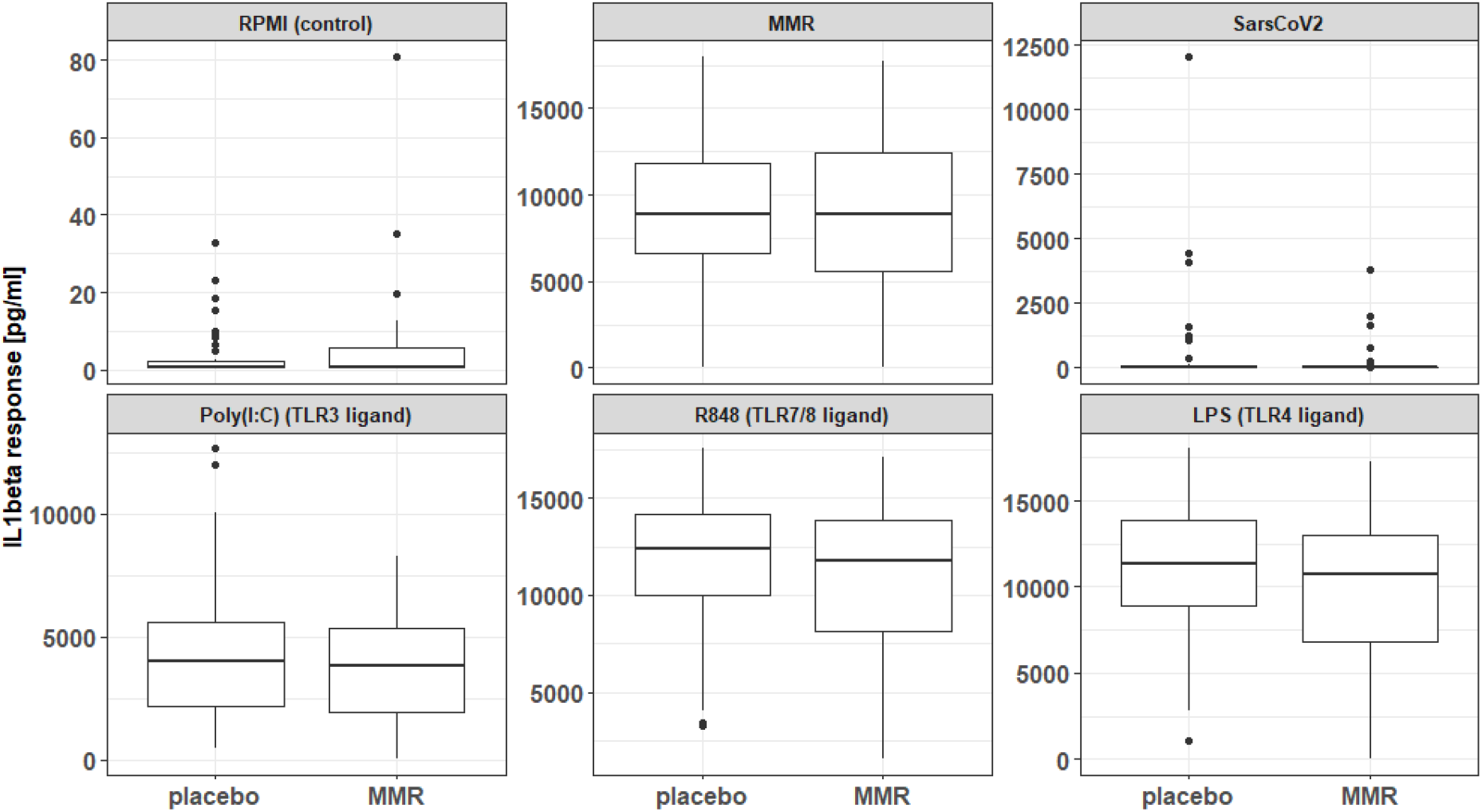
IL-1β responses to heterologous stimulation

**Figure 4:**
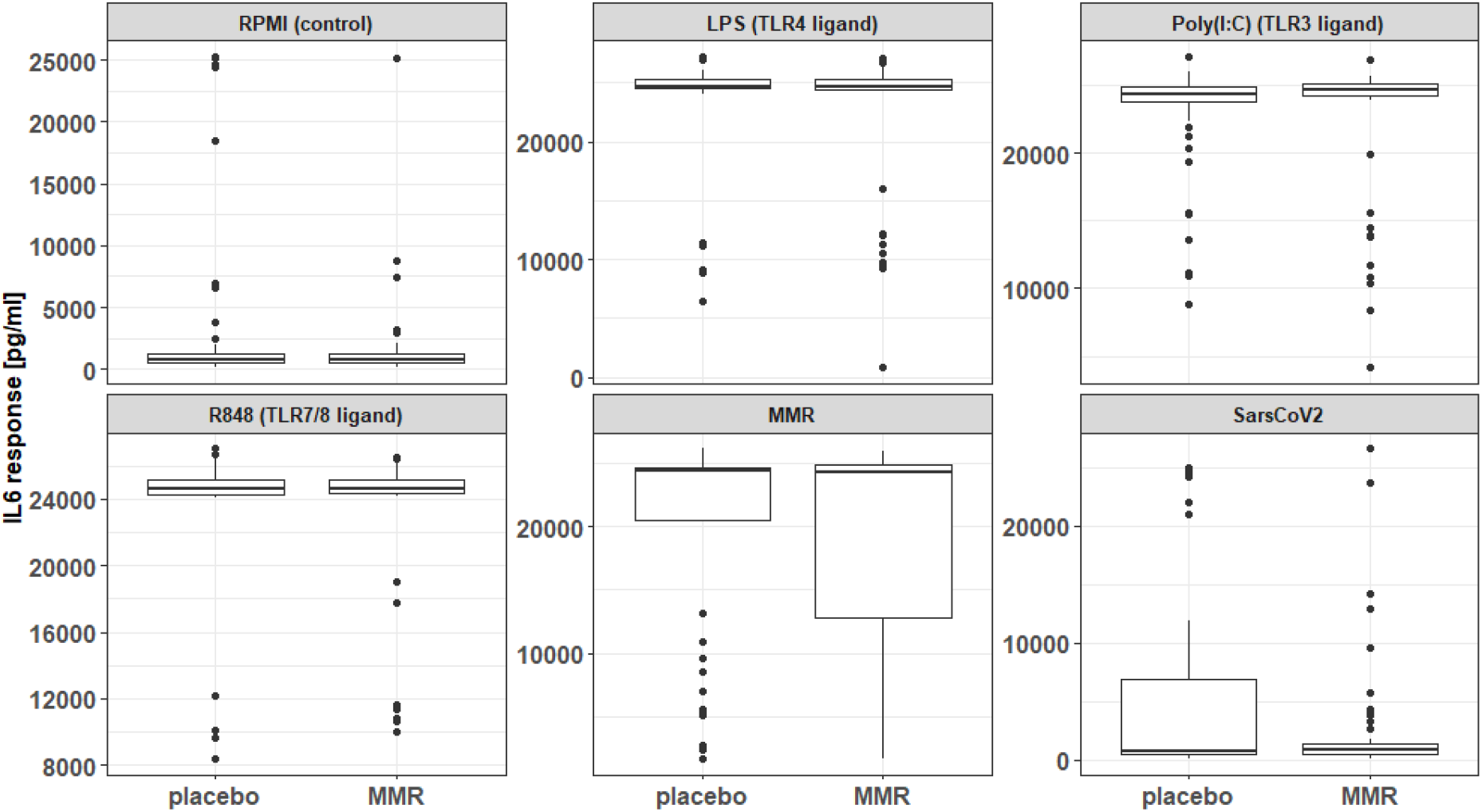
IL-6 responses to heterologous stimulation

**Figure 5:**
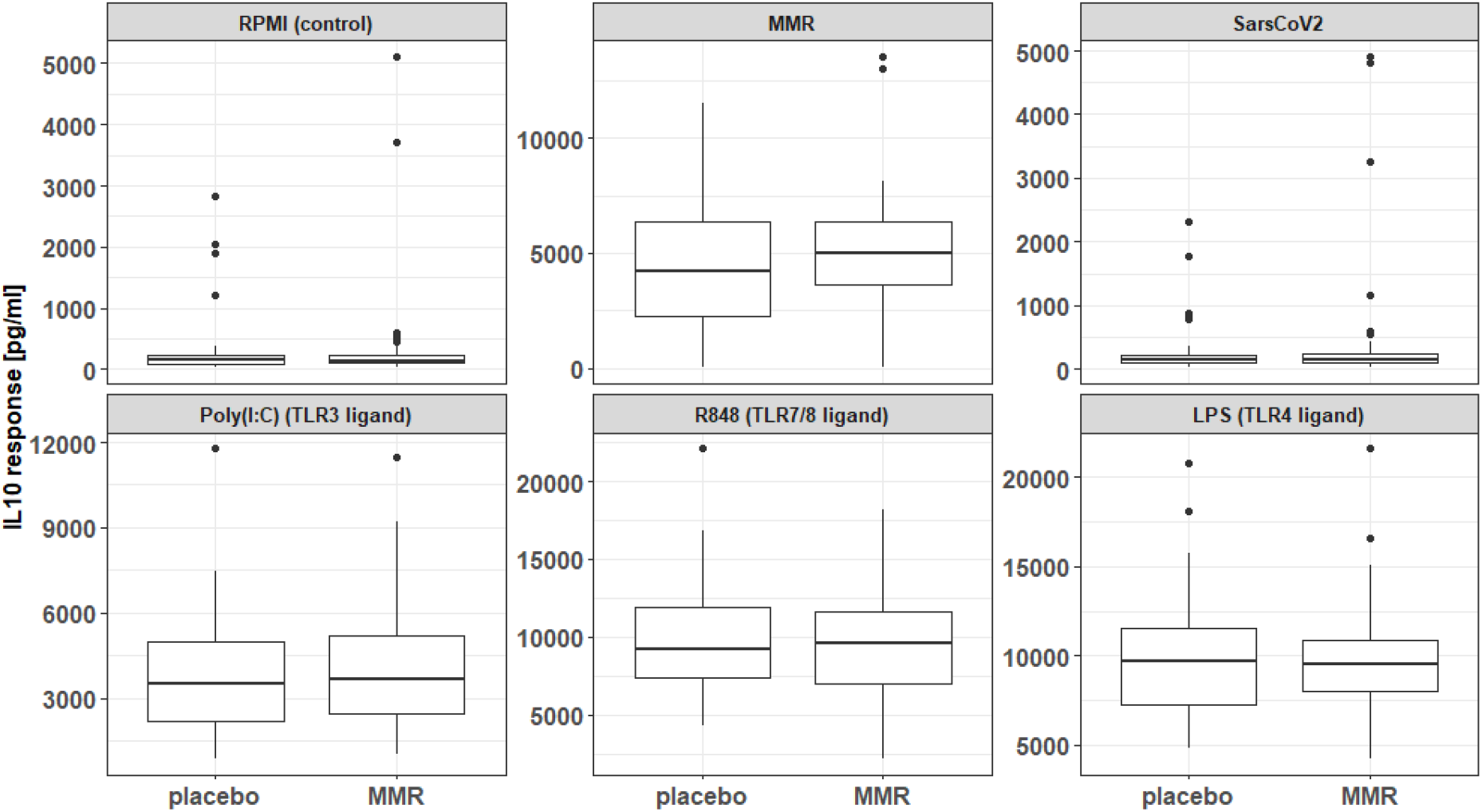
IL-10 responses to heterologous stimulation

**Figure 6:**
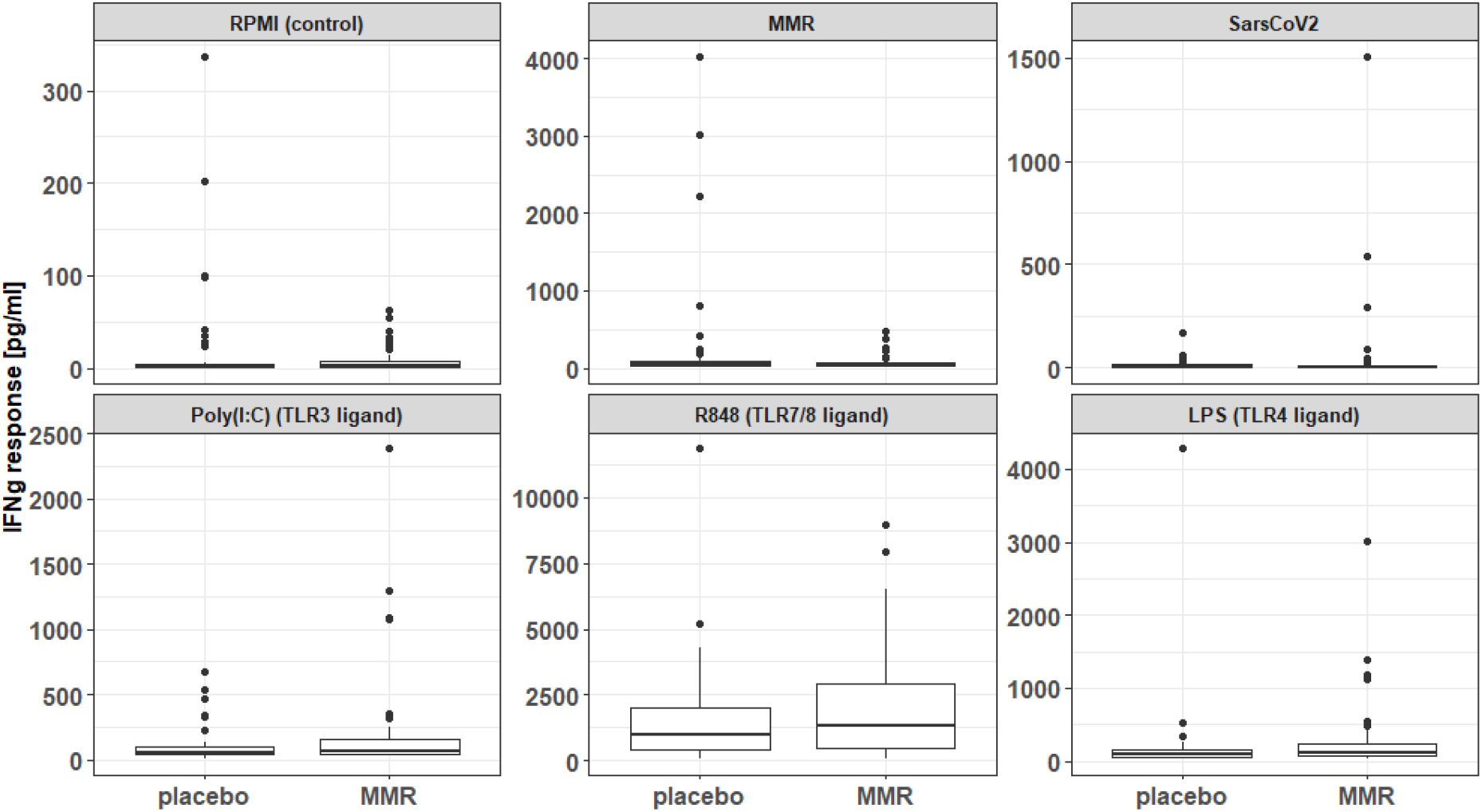
IFN-γ responses to heterologous stimulation

### Effects of MMR vaccination on serological responses

In the SARS-CoV-2 neutralization assay, the unadjusted geometric mean (95% CI) IC50 in the MMR group was 507.6 (385.9; 656.1) and in the placebo group was 515.7 (415.0; 638.6); ratio of geometric means (95% CI) 1.0 (0.7; 1.5) adjusted for sex and quarter of the year; Figure 7.

**Figure 7:**
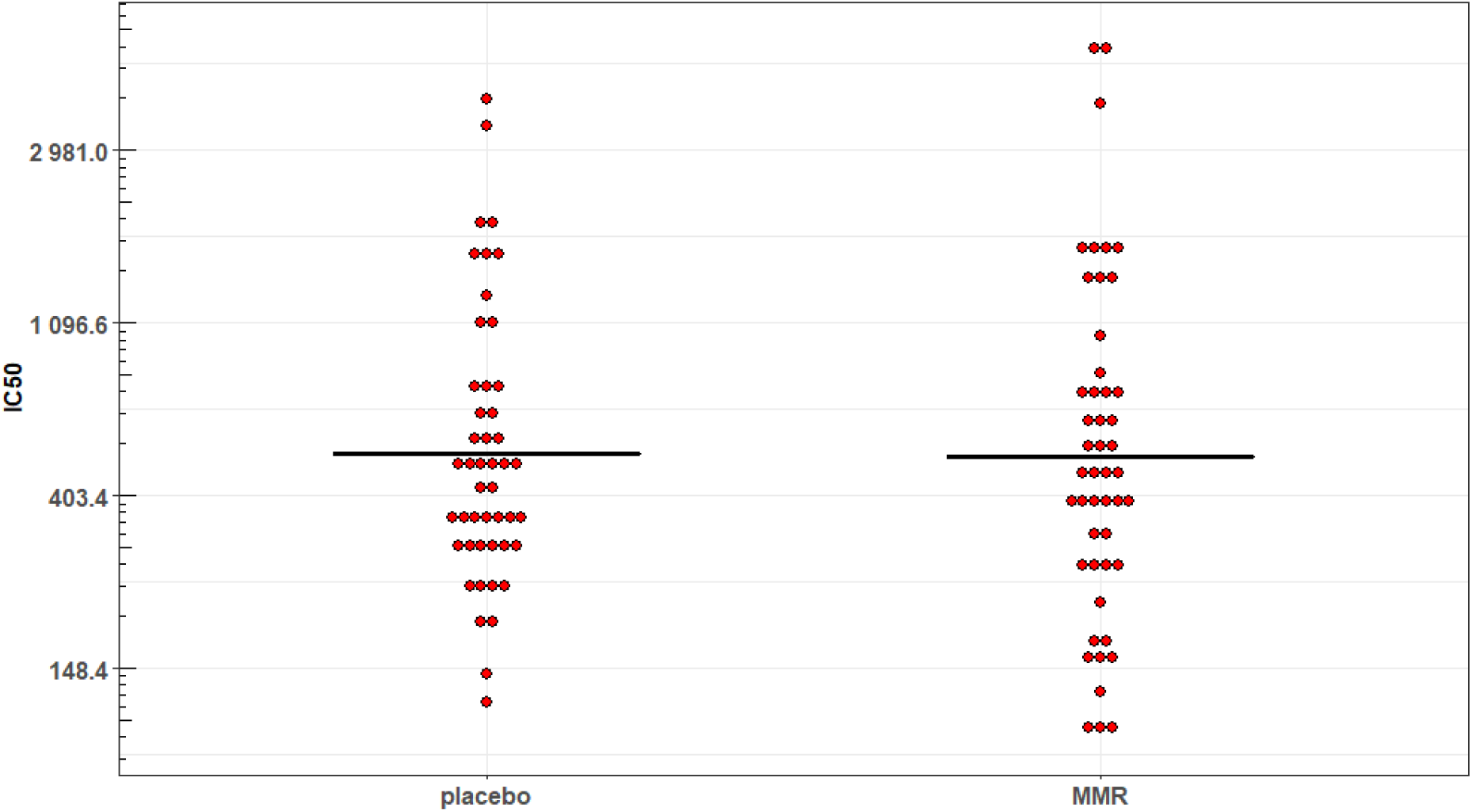
Neutralization assay (IC50)

Measles IgG titres were tested with a qualitative assay for the first 12 participants and with a quantitative assay for the subsequent 83 participants. Forty-five of 48 (94%) participants in the MMR group had a measles IgG titre ≥ 16.5 AU/ml, consistent with immunity. Thirty-nine of 47 (83%) participants in the placebo group had a measles IgG titre ≥ 16.5 AU/ml, consistent with immunity. Those in the MMR group had 3.1 times the odds of being classified immune to measles (measles IgG titres ≥ 16.5 AU/ml) compared to those in the placebo group (OR 95% CI 0.8; 14.8), n = 95.

Sensitivity analysis excluding the six participants who developed COVID-19 prior to their blood draw did not change the findings of the analyses of the chemokine and cytokine responses, the IC50 results or the measles IgG results.

## Discussion

In this relatively young (three quarters of participants under the age of 50) and largely female (two thirds of participants identified as female) study population, pre-exposure to MMR vaccine was generally not associated with changes in cytokine and chemokine responses of stimulated PBMCs 15 weeks after SARS-CoV-2 mRNA vaccination. MMR vaccination was associated with a signifoicant increase of TNF-α production in response to an additional ex-vivo stimulation with the MMR vaccine. The SARS-CoV-2 neutralization IC50 values did not differ significantly between MMR and placebo groups.

The overwhelming majority of TNF-α comes from monocytes and macrophages (Vassalli, 1992). T-cells can produce small amounts of TNF-α, but much lower than myeloid cells. While IFN-γ can amplify TNF-α production by monocytes and macrophages, we did not observe increased IFNg production in response to MMR stimulation. IFNg is produced mostly by T-cells, and partially from NK cells. The difference in TNF-α production in response to MMR stimulation may represent differences in gene transcription mediated by epigenetic changes (Saeed et al., 2014). If our observation is reproducible, this mechanism would need exploration in future. However, it is possible that the difference we observe is simply a spurious finding. A significant proportion (83%) of participants in the placebo group had measles IgG titres ≥ 16.5 AU/ml, consistent with immunity. On the other hand, the measles IgG titres in 6% of participants in the MMR group were < 16.5 AU/ml, not consistent with immunity. Nonetheless, the MMR group tended towards greater odds of being considered immune to measles (OR 3.1; 95% CI 0.8; 14.8).

The observed results from our PBMC stimulation study does not support our hypothesis that pre-exposure to MMR vaccination induces lasting increased sensitivity of innate immune cells to subsequent heterologous stimuli. The innate immune cells circulating at the time of MMR / placebo vaccination are not expected to remain present 4-5 months down the line. But prior work on trained immunity demonstrate induced memory response in bone marrow progenitors and long lasting tissue resident innate immune cells, although this needs to be formally demonstrated for MMR vaccination. Furthermore, preexposure to MMR had no notable effect on the neutralizing antibody response to subsequent SARS-CoV-2 mRNA vaccination.

The difference in IL-6 responses between treatment groups could not be assessed as the IL-6 response to all TLR ligands and to repeated MMR exposure were truncated by the detection ceiling of the assay. This is a key limitation of our study as secretory IL-6 levels are a hallmark of immune tolerance and training. The absence of repeated measures in this sub-study and the variability in the exposure-sampling window further limits the strength of our findings. However, the random treatment assignment of participants addresses concerns about confounding introduced by variability in the exposure-sampling window.

Our inability to demonstrate a lasting trained immune response supports the observed absence of clinical protection by a measles-containing vaccine against COVID-19 in our own clinical trial. An unanswered question remains whether a notable trained immune response existed after MMR exposure which was subsequently negated by exposure to the mRNA SARS-CoV-2 vaccine. Reversal of trained immunity responses and protection by other live attenuated vaccines by inactivated vaccines has been previously shown in case by BCG vaccination followed by DTP (Aaby et al., 2007). Whether a similar inhibitory effect in MMR vaccinated individuals was induced by the mRNA COVID19 vaccine is not possible to assess in the current study. While our sub-study data cannot speak directly to such vaccine-vaccine immunological interaction, the absence of interaction on clinical outcomes in the larger CROWN CORONATION study population suggests this was not the case. While a positive priming response of MMR prior to mRNA SARS-CoV-2 vaccination would have been desirable, it is a solace that we observed no deleterious vaccine-vaccine immunological interaction either in the PBMC stimulation responses or the SARS-CoV-2 neutralization assay.

## Supporting information

CROWN CORONATION Investigators

## Data Availability

All data produced in the present study are available upon reasonable request to the authors.

## Acknowledgements

Special thanks to our colleagues at the COVID-19 Therapeutics Accelerator (Gail Rodgers, Scott Miller, Janice Culpepper, Keith Klugman), who worked closely with the investigators on the CROWN CORONATION trial.

