## Supplementary material for "THE EFFECT OF THE MEASLES, MUMPS AND RUBELLA VACCINE ON INNATE AND ADAPTIVE IMMUNE RESPONSES IN PERSONS RECEIVING A SARS-COV-2 mRNA VACCINE": CROWN CORONATION Investigators

* Principal investigator


**Washington University School of Medicine, St. Louis, Missouri, United State of America**

Michael S. Avidan*

Shabaana Khader

Erik Dubberke*

Mary Politi

Linda Yun

Leon du Toit

Christian Guay

Anne K. DeSchryver

Sherry McKinnon

Benjamin Swan

Ananya Gupta

**Centre for Perioperative Medicine, Department of Targeted Intervention, University College London, United Kingdom**

S. Ramani Moonesinghe*

Laurence Lovat*

Dermot McGuckin

**Comprehensive Clinical Trials Unit, University College London, United Kingdom**

Hakim-Moulay Dehbi (Lead statistician)

Emilia Caverly

Gemma Jones

Nicholas Freemantle

**Department of Internal Medicine, Radboud University Medical Center, Nijmegen, The Netherlands**

Mihai G. Netea

**Medinel, Western Cape, South Africa**

Annalene Nel

**Department of Anaesthesia and Perioperative Medicine, Groote Schuur Hospital and University of Cape Town, South Africa**

Bruce Biccard*

Margot Flint

Malcolm Miller

Leon du Toit

**Division of Emergency Medicine, Groote Schuur Hospital, Cape Town, South Africa**

Willem Stassen

**Desmond Tutu Health Foundation, Cape Town, South Africa**

Linda-Gail Bekker*

Yashna Singh

Doerieyah Reynolds

Catherine Orrell

**Desmond Tutu HIV Centre, Institute of Infectious Disease and Molecular Medicine, University of Cape Town, South Africa**

Katherine Gill*

Mellissa LeFevre

Menna Duyver

Gina Itzikowitz

**Wits RHI, University of the Witwatersrand, Johannesburg, South Africa**

Sinead Delany-Moretlwe*

S. Helen Rees*

Darshnika Lakhoo

Edwin Mkhwanazi

Miliswa Magongo

**University of Witwatersrand, Clinical HIV Research Unit, Helen Joseph Hospital, Johannesburg, South Africa**

Noluthando Mwelase*

Thembi Masiye

Nokuphiwa Mbhele

Jessica Maritz

**Africa Health Research Institute (AHRI), Durban, South Africa**

Limakatso Lebina*

**Perinatal HIV Research Unit, University of the Witwatersrand, Johannesburg, South Africa**

Limakatso Lebina*

Neil A. Martinson

Leisha Genade

Lesego Mmolawa

**Setshaba Research Centre, Pretoria, South Africa**

Khatija K. Ahmed*

Kgaogelo Molapo

Dr Zinhle Zwane

Sr Thando Moloelang

**The Aurum Institute, Johannesburg, South Africa**

Kathy T Mngadi*

Sibekezelo Msomi

Mashudu Mahlangu

Yajna Duki

Anja N Henning

**HIV Prevention Research Unit, South African Medical Research Council, Durban, South Africa**

Villeshni Asari*

Reshmi Dassaye*

**Department of Anaesthesiology, University of the Free State, Bloemfontein, South Africa**

Edwin Turton*

**JOSHA Research, Bloemfontein, South Africa**

Edwin Turton*

Ntaoleng Frangenie Coangae

**Family Centre for Research with UBUNTU (FAMCRU), Stellenbosch University, Tygerberg, South Africa**

Samantha H Fry*

Jeannine Du Bois

Yasmeen Akhalwaya

Phoebe Ganjana

**Noguchi Memorial Institute for Medical Research, University of Ghana, Accra, Ghana**

George Kyei*

Kwadwo Koram*

Kwadwo Kusi

Susan Adu- Amankwah

**Centre for Infectious Diseases Research in Zambia (CIDRZ), Lusaka, Zambia**

Izukanji Sikazwe*

Chikumbutso Chipeta

Joyce Chinyama Chilekwa
